# Sit-to-Stand Adaptations in Pre-Operative Cam-Type Femoroacetabular Impingement: A Kinematic and Coordination Analysis

**DOI:** 10.64898/2026.10.26.26366097

**Authors:** Patrick Ippersiel, Andrew Tucker, Ariane Parisien, George Grammatopolous, Andrew D. Speirs, Mario Lamontagne, Paul E. Beaulé

## Abstract

**Background:** Individuals with cam-type femoroacetabular impingement (FAI) demonstrate altered lower-limb biomechanics during functional activities, yet little is known about inter-joint coordination during sit-to-stand (STS). This study compared sagittal-plane kinematics, inter-joint coordination, and coordination variability during STS in individuals with and without cam-type FAI.

**Methods:** Twenty-four adults with symptomatic cam-type FAI and 19 healthy controls completed five STS trials while three-dimensional motion capture and ground reaction force data were collected. Sagittal-plane hip, knee, ankle, pelvis, and spine kinematics were analyzed using statistical parametric mapping and discrete measures. Spine-hip and hip-knee coordination variability were quantified using modified vector coding. Group differences were evaluated using independent t-tests and mixed ANOVAs.

**Results:** Participants with FAI demonstrated reduced early-phase hip range of motion and greater peak knee flexion during STS compared with controls. Statistical parametric mapping did not reveal statistical group differences in single joint kinematics during early and late phase STS. A significant group-by-phase interaction was observed for spine-hip coordination variability, with individuals with FAI exhibiting greater variability during early STS (p = 0.010, d = 0.92), but not during late STS. No between-group differences were identified for hip-knee coordination variability.

**Conclusions:** Individuals with cam-type FAI demonstrate altered spine-hip movement organization during STS, characterized by greater early-phase coordination variability and reduced hip flexion range of motion.

**Clinical significance:** These findings suggest that movement strategies in people with FAI during STS may be more strongly reflected in variability of inter-joint coordination patterns than single joint kinematics.

## 1. INTRODUCTION

Femoroacetabular impingement (FAI) is a well-established cause of hip pain among active young adults (Beaulé et al., 2007; Wisniewski and Grogg, 2006) and has been linked to the development of early-onset osteoarthritis (Agricola et al., 2024, 2013; Ganz et al., 2003). Cam-type, the most common morphologic subtype of FAI, is characterized by an aspherical head-neck junction that can produce abnormal femoroacetabular contact during terminal hip motion, such as those involving flexion, adduction, and internal rotation (Ganz et al., 2003; Griffin et al., 2016). Functional tasks that load the hip in or near these positions can provoke symptoms and are associated with altered hip, pelvis, and lower-extremity movement patterns in individuals with cam-type FAI (Samaan et al., 2017; Spiker et al., 2022; Zhang et al., 2025). These adaptations may arise partly in response to the interaction between cam morphology and acetabular or spino-pelvic anatomy (Grammatopoulos et al., 2018) and may reflect altered femoropelvic coordination or joint loading strategies which could contribute to secondary cartilage or labral stress (Diamond et al., 2017; Kumar et al., 2014).

Prior biomechanical research in individuals with cam-type FAI has commonly focused on gait and squatting, with evidence of reduced peak hip extension and sagittal-plane hip range of motion during gait, as well as altered hip–pelvis mechanics and reduced sagittal-plane pelvic motion during squatting (Bagwell et al., 2016; Lamontagne et al., 2009; Yarwood et al., 2022). Further, following corrective surgery, FAI patients have shown greater squat depth which was accompanied by increases in knee flexion and ankle dorsiflexion, suggesting that performance may be modified through redistribution of movement across the lower-extremity kinetic chain (Lamontagne et al., 2011). In contrast, the sit-to-stand (STS) task has received comparatively less attention despite being a high-frequency functional movement that imposes substantial biomechanical demands on the lower limbs (Dall and Kerr, 2010; Janssen et al., 2002). Existing STS work suggests that individuals with FAI demonstrate altered functional performance, lower-extremity moment contributions, and muscle activity during this task (Samaan et al., 2017; Spiker et al., 2022). However, these studies have emphasized discrete biomechanical measures, which may not fully capture the coordinated trunk, hip, knee, and ankle motions required to transfer body mass from a seated to standing position. Whether individuals with cam-type FAI show changes in inter-joint coordination and/or coordination variability during STS remains unclear.

Inter-joint coordination and variability provide insight into neuromuscular control strategies beyond discrete kinematic measures. Modified vector coding quantifies dynamic relationships between joint motions, yielding coupling angles and coupling angle variability that describe both the organization and consistency of movement across repeated trials (Needham et al., 2014, 2015). This is clinically relevant because reduced coordination variability has been interpreted as a less adaptable movement strategy that may concentrate repetitive loading on a smaller region of tissue (Hamill et al., 2012, 1999). Given the proposed role of altered hip mechanics and joint loading in cam-type FAI, applying this approach to STS may reveal subtle differences in movement strategy that are not evident from sagittal-plane kinematics alone.

The purpose of this study was to estimate the effect to which sagittal plane kinematics, inter-joint coordination, and coordination variability during STS differ in people with and without FAI. We hypothesized that cam-type FAI patients would show reduced hip joint ROM and altered spinal kinematics and coordination variability, compared to healthy controls.

## 2. METHODS

### 2.1. Overview

This cross-sectional study was completed at the Human Movement Biomechanics Laboratory, at the University of Ottawa (Ontario, Canada). Each participant underwent biomechanical testing and completed questionnaires in a single testing session. Institutional ethics approvals were obtained prior to study initiation (Ottawa Health Science Network and uOttawa – Research Ethics Board Number: 20190456-01H and H-09-19-4994 - OTH-4994 respectively). All study participants provided informed consent.

### 2.2 Participants

Participants were recruited from a hip preservation practice with two orthopedic surgeons through various sources (e.g., orthopedic clinic referrals, pre-operative screening, or departmental research registry). Individuals in the cam-type FAI cohort were skeletally mature adults (>18 years) scheduled for arthroscopic cam resection, who reported localized groin or lateral hip pain for longer than six months and demonstrated a positive impingement sign. Diagnosis of cam-type FAI was confirmed through clinical examination and radiographic evaluation. Radiographic criteria included a Tönnis grade of 0 - 1, absence of acetabular dysplasia (lateral center-edge angle >25°), absence of acetabular overcoverage (LCE >39°), and an α-angle greater than 55° on multiplanar imaging. In the event patients had bilateral cam-type impingement, only the pre-surgical, symptomatic limb was used for our analyses.

Healthy control participants were required to be over 18 years of age, demonstrate a normal femoral head-neck contour with no evidence of dysplasia, and be willing to complete all required assessments. Participants were excluded if they had a prior joint replacement in any lower-limb joint, a lower-limb or lumbar spine injury within the previous year, metal prostheses, BMI greater than 30 kg/m² or excessive waist circumference (>102cm in men and >88cm in women), a strong family history of early osteoarthritis, or other serious medical conditions.

### 2.3. Demographics and questionnaires

The following demographic data were collected: age, biological sex, height, and weight. General hip symptoms and functional limitations were evaluated using the Hip disability and Osteoarthritis Outcome Score (HOOS), which assesses self-report pain, symptoms, activities of daily living, sport and recreation, and hip-related quality of life (Nilsdotter et al., 2003). The HOOS is a well-validated patient-reported outcome measure with established reliability, validity, and responsiveness in populations with hip pathology, and can discriminate between individuals with hip disorders and healthy controls (Kemp et al., 2013).

### 2.4. Protocol

This manuscript describes one task (STS) from a broader biomechanical data collection where participants with and without FAI completed a battery of functional tasks. For the standardized STS task, each participant began seated upright on an adjustable bench-mounted force plate. The seat height was individually adjusted to achieve approximately 90° of knee flexion to control for differences in participant anthropometrics. Each foot was positioned at the center of two separate floor-mounted force plates (Figure 1). Data were collected from the FAI group prior to their surgery.

**Figure 1.** Standardized sit-to-stand protocol. Representative participant shown in the initial seated position (left) and at upright standing (right). The participant was seated on a bench-mounted force plate with each foot positioned on a separate floor-mounted force plate and completed the task while maintaining a standardized upper-extremity position.

Participants were asked to flex their shoulders to 90° in the scapular plane and flex their elbows to 90°. From this starting position, participants were instructed to rise to full knee extension at a self-selected speed while maintaining the initial arm position. Upon reaching full extension, they held the standing posture for 5 seconds before returning to the seated position on the bench to reset. Five consecutive trials were performed under these conditions without a prescribed rest interval. To ensure consistency, participants were instructed not to use their upper extremities for momentum generation. Foot placement was standardized to shoulder-width apart and verified prior to each trial. All participants wore comfortable, closed-toe footwear and performed the task unassisted.

### 2.5. Data Acquisition and Processing

Three-dimensional kinematic data were collected at 200 Hz with 45 reflective markers using a 10-camera system (Vicon MX, UK). Markers were placed symmetrically across the head, trunk, pelvis, upper and lower extremities based on the University of Ottawa Motion Analysis Model marker set (Mantovani and Lamontagne, 2017). Ground reaction forces (GRFs) were recorded at 1,000 Hz using force plates (FP4060-08, Bertec Corporation, Columbus, USA) positioned under each foot and a bench-mounted force plate (Model 9262BA, Kistler Group, Winterthur, Switzerland) on the seat. A static standing calibration trial was conducted prior to dynamic testing to define participant joint centers and segment coordinate systems. Data were synchronized and processed in Nexus software using the University of Ottawa Motion Analysis Model (UOMAM). Specifically, data were labeled, gap filled, and filtered (zero-lag, fourth order Butterworth filter, cut-off frequency = 6 Hz). Lower limb 3-D motions were calculated according to established methods (Davis III et al., 1991; Kadaba et al., 1990). Joint angles were calculated based on a YXZ Cardan sequence, and pelvic angles were measured relative to a global coordinate system. Joint and segment angles were zeroed based on each participant’s neutral position as determined by a standing static trial. Trials with extensive marker loss or unrecoverable data gaps during processing were excluded. Data were then exported to MATLAB (R2023b) using custom scripts for further processing and analyses.

### 2.6. Event identification

The STS task was partitioned into early and late phases using a custom MATLAB script. The early phase was defined from movement onset to seat-off, and the late phase from seat-off to movement end. Seat-off was detected from unloading of the bench vertical ground reaction force, defined as the first sustained point at which bench load fell below 5% of the initial seated baseline load (Ippersiel et al., 2018). Movement onset and movement end were automatically identified from a composite sagittal-plane movement velocity signal derived from available trunk, spine, hip, and knee angle trajectories and pre-determined threshold values. Event-detection plots for each trial were visually inspected by an experienced researcher and manually adjusted when necessary. After identifying key movement events, kinematic data were segmented into early-phase, late-phase, and full-cycle intervals, and each interval was time-normalized to 100% of the STS phase.

### 2.7. Outcome Measures

#### 2.7.1. Joint angles and range of motion

Kinematic outcomes included peak joint angles and ranges of motion for the hips, knees, ankles, pelvis, and spine in the sagittal plane. For FAI patients, data were extracted for their affected limb. For healthy controls, data were extracted for their limb (right vs. left) based on a randomization process. For each participant, data were averaged across all available trials, for the early phase, late phase, and full cycle.

#### 2.7.2. Vector coding analyses (coordination and coordination variability)

Sagittal plane hip-knee and spine-hip inter-joint coupling angles (CA) and coupling angle variability (CAV) were calculated using modified vector coding for the STS task (Chang et al., 2008; Ippersiel et al., 2024; Needham et al., 2014). For the hip-knee pairing, the proximal and distal segments were defined as hip and knee, respectively. For the spine-hip pairing, the proximal and distal segments were defined as spine and hip, respectively. Distal and proximal sagittal plane joint angle waveforms were plotted on the vertical and horizontal axes of an angle-angle plot.

Vector coding was performed separately for time-normalized early and late phases of STS. CAs were calculated from the vector connecting consecutive points of the angle-angle plot and expressed from 0° and 360° and categorized into four coordination patterns: in-phase distal dominant, in-phase proximal dominant, anti-phase distal dominant, and anti-phase proximal dominant (Figure 2, Supplemental Figure 1). In-phase indicates that two joints are rotating in the same direction, while anti-phase indicates rotations in opposite directions. Proximal or distal dominant coordination reflects that the angular change of one joint was greater than that of the other one over a set time interval.

**Figure 2.**
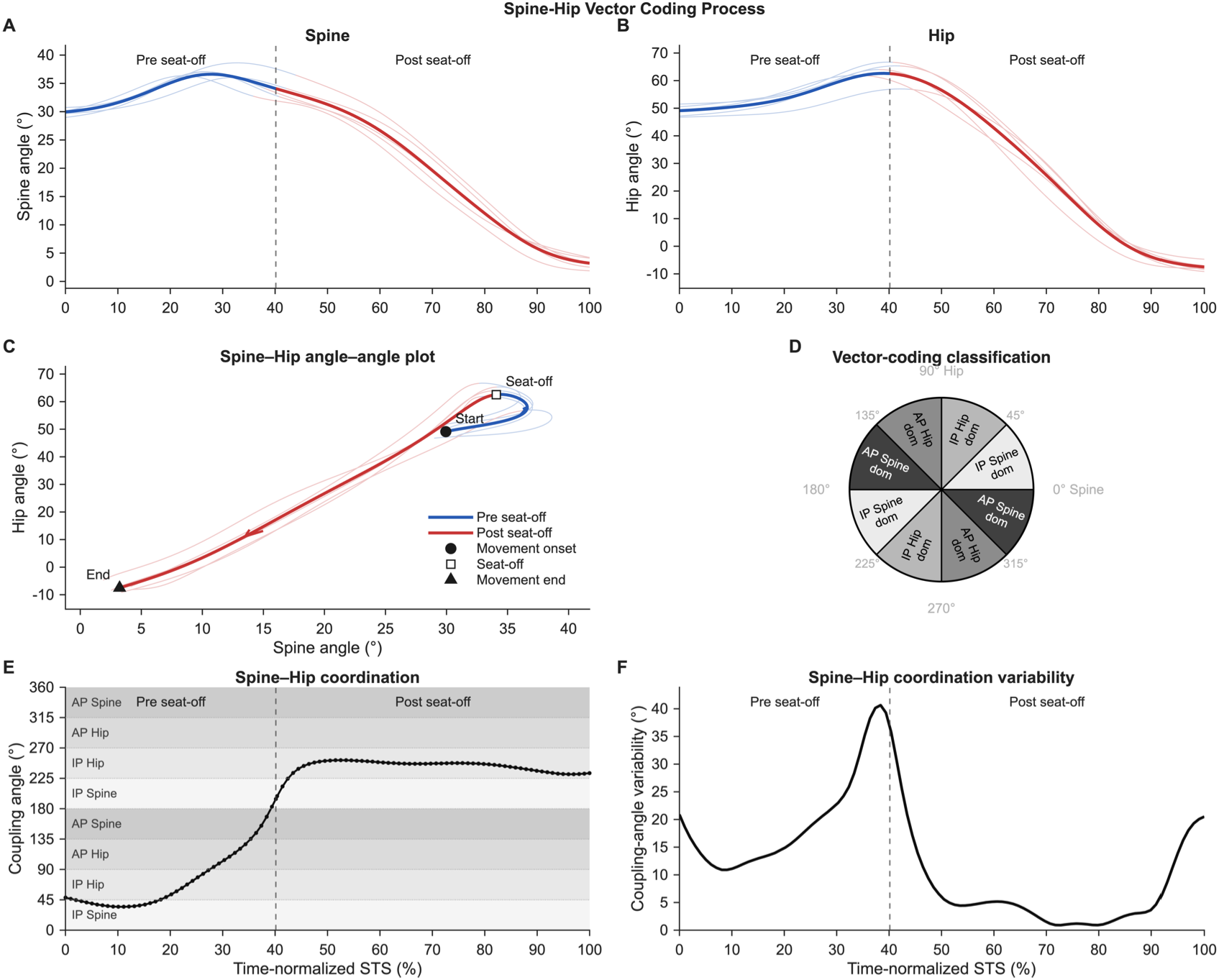
Illustrative spine–hip vector-coding analysis for a representative participant with FAI. Panels show spine and hip waveforms, the angle–angle trajectory, coordination-pattern classification, coupling angle, and coupling-angle variability across STS. The full-task representation is shown for illustrative purposes only; primary analyses were performed separately for the independently time-normalized early and late phases

Coordination pattern frequencies (i.e., bin frequencies) were calculated at the trial level and then averaged within participants to preserve trial-level information prior to group summaries (Napoli et al., 2025). Mean CAs were calculated using circular statistics. Coordination variability was quantified as coupling angle variability (CAV). For each participant, pointwise CAV was calculated across repeated trials using circular dispersion of CAs. Early-and late-STS CAV was then calculated by averaging the pointwise CAV waveform across individual phases. Simply, pointwise CAV represents how consistent or variable the coordination pattern was across repeated trials at each moment of the STS phase.

### 2.8. Statistical Analysis

Descriptive statistics were calculated for all demographic factors, patient questionnaires, and outcome variables.

To examine between-group differences in sagittal plane kinematics, statistical parametric mapping (SPM) was applied across the time-normalized early and late STS phases, separately for the hip, knee, ankle, spine, and pelvis (Pataky, 2010). Two-tailed independent sample t-tests (spm1d.stats.ttest2) were used. To account for potential between-participant differences in initial joint or segment position, each waveform was offset-referenced by subtracting the mean value of the first five samples from the entire waveform prior to analysis. Thus, SPM analyses evaluated between-group differences in angular change from the initial seated position rather than absolute joint or segment angle. As a secondary analysis, Welch’s t-tests were run to compare FAI and healthy groups on discrete biomechanical measures, namely peak angle and joint range of motion for the hip, knee, ankle, spine, and pelvis, separately for the early, late, and full STS phases. Hedges g quantified effect sizes.

To examine group differences in spine-hip and hip-knee coordination variability (CAV), two-way mixed ANOVAs were used. This test examined the effects of group (FAI vs. Healthy) and their interaction (group x phase), separately for the spine-hip and hip-knee joint pairs. When a statistically significant interaction occurred, post-hoc Bonferroni-adjusted pairwise comparisons were performed. Effect sizes were reported using ηₚ² and Cohen’s d.

Coordination-pattern frequencies were summarized descriptively because the four categories form interdependent proportions of the same movement phase and summed to 100%. Therefore, inferential testing was not considered appropriate.

Assumptions for statistical tests were verified. Normality was assessed using the Shapiro-Wilk test and inspection of Q-Q plots, and homogeneity of variance was examined via Levene’s test. Statistical significance was set at p < 0.05 for all tests. Statistical analyses were conducted using Jamovi (V.2.7.7.0) and MATLAB.

### 2.9. Sample size

A retrospective sample-size calculation was performed using a previously reported between-group difference in hip-lumbar coordination variability during STS (Ippersiel et al., 2018). Using *α* = 0.05. 80% power, and an estimated effect size of Hedges g = 0.86, approximately 22 participants per group would be required. Analyses were performed in G*Power 3.1.

## 3. RESULTS

### 3.1. Data and participant characteristics

Data from three participants (2 FAI and 1 healthy) were excluded due to technical problems during data collections. Biomechanical data were screened for statistical outliers which resulted in the omission of 47/1648 waveforms (2.85%). The final sample comprised 24 participants with FAI and 19 healthy controls. Of the 24 FAI patients, 9 had unilateral cam-type FAI while 15 had a bilateral diagnosis. In all cases, the symptomatic, pre-surgical limb was used for analyses. Based on Welch’s t-tests, participants with FAI were statistically older than healthy controls and had smaller scores on all HOOS subscales (p < 0.05). The FAI group had a descriptively higher proportion of men than the control group (95.8% vs. 73.7%); however, this was not statistically different (p > 0.05). Full participant characteristics are listed in Table 1.

**Table 1.** Participant Characteristics.

| Characteristic | FAIS | CTRL |
| --- | --- | --- |
| Participants, n | 24 | 19 |
| Age, years | **33.2 (9.3)** | **26.4 (5.2)** |
| Female sex, n (%) | 1 (4.2%) | 5 (26.3%) |
| Height, cm | 180.9 (5.9) | 176.6 (10.2) |
| Weight, kg | 82.6 (10.4) | 78.0 (12.2) |
| BMI, kg/m<sup>2</sup> | 25.2 (2.7) | 25.0 (3.3) |
| HOOS Symptoms | **49.4 (15.1)** | **96.4 (7.0)** |
| HOOS Pain | **57.0 (20.6)** | **99.9 (0.6)** |
| HOOS Activities of Daily Living | **67.5 (17.9)** | **99.9 (0.4)** |
| HOOS Sport/Recreation | **45.6 (24.6)** | **100.0 (0.0)** |
| HOOS Quality of Life | **30.8 (18.2)** | **100.0 (0.0)** |

*Values are mean (SD) unless otherwise indicated. Sex is reported as female n (%). HOOS = Hip disability and Osteoarthritis Outcome Score; scores range from 0 to 100, with higher scores indicating better status. FAIS = Femoroacetabular impingement syndrome. CTRL = Healthy controls. Bolded values denote statistical differences ( $p < 0.05$ ).*

### 3.2. Group differences in sagittal plane kinematics

Statistical parametric mapping revealed no significant group differences in sagittal plane hip, knee, ankle, pelvis, and spine angles for the early or late phases of the STS task (p > 0.05, Figure 3, Supplemental Figure 2). Trends were observed and suggested decreased hip flexion and pelvic anterior tilting, alongside greater spinal flexion in FAI patients during the latter portion of early STS (i.e., close to seat-off); however, these were not statistically significant.

**Figure 3.**
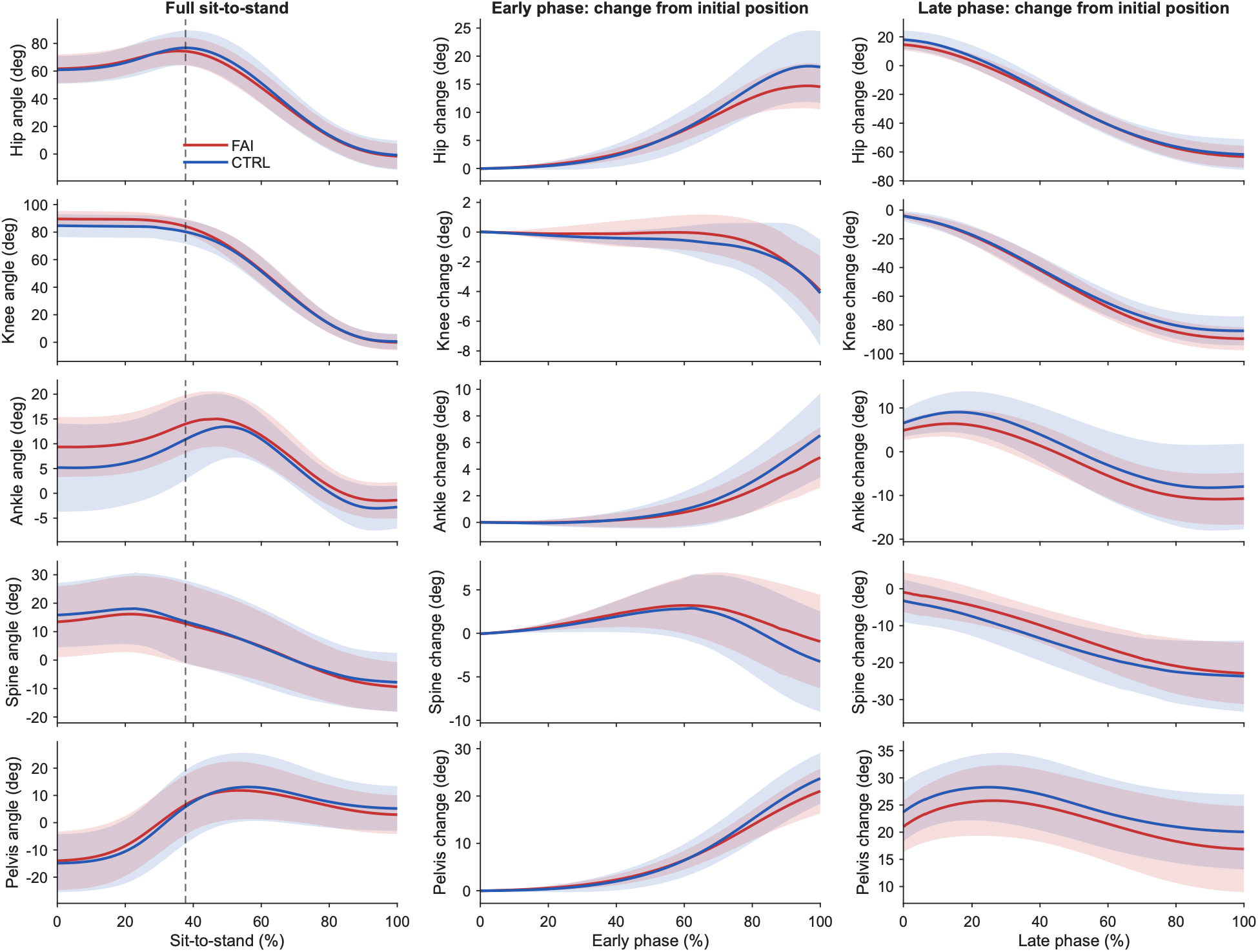
Sagittal-plane kinematic waveforms during sit-to-stand in individuals with femoroacetabular impingement (FAI) and Healthy controls (CTRL). Group mean waveforms are shown for the FAI affected limb/segment and CTRL limb. The left column presents absolute joint/segment angles across the full sit-to-stand task, while the middle and right columns present offset-referenced angular change from the initial seated position during the early and late phases, respectively. Shaded colored bands represent ±1 SD. Grey vertical shading indicates regions of statistically significant between-group differences identified using statistical parametric mapping. Positive values indicate greater flexion and anterior tilt (pelvis).

Results of Welch’s t-tests reporting group differences in ROM and peak angles by joint and phase are listed in Table 2. Of note, compared to healthy controls, FAI patients had less hip ROM during the early phase (mean difference = −3.50°; 95% CI: −6.85 to - 0.15) and greater knee ROM during the late phase (mean difference = 5.60°; 95% CI: 0.04 to 11.16). Otherwise, greater peak knee flexion was observed during early-phase STS (mean difference = 4.90°; 95% CI: 0.54 to 9.26), late-phase STS (mean difference = 5.10°; 95% CI: 0.75 to 9.45), and the full task (mean difference = 4.90°; 95% CI: 0.54 to 9.26).

**Table 2.**
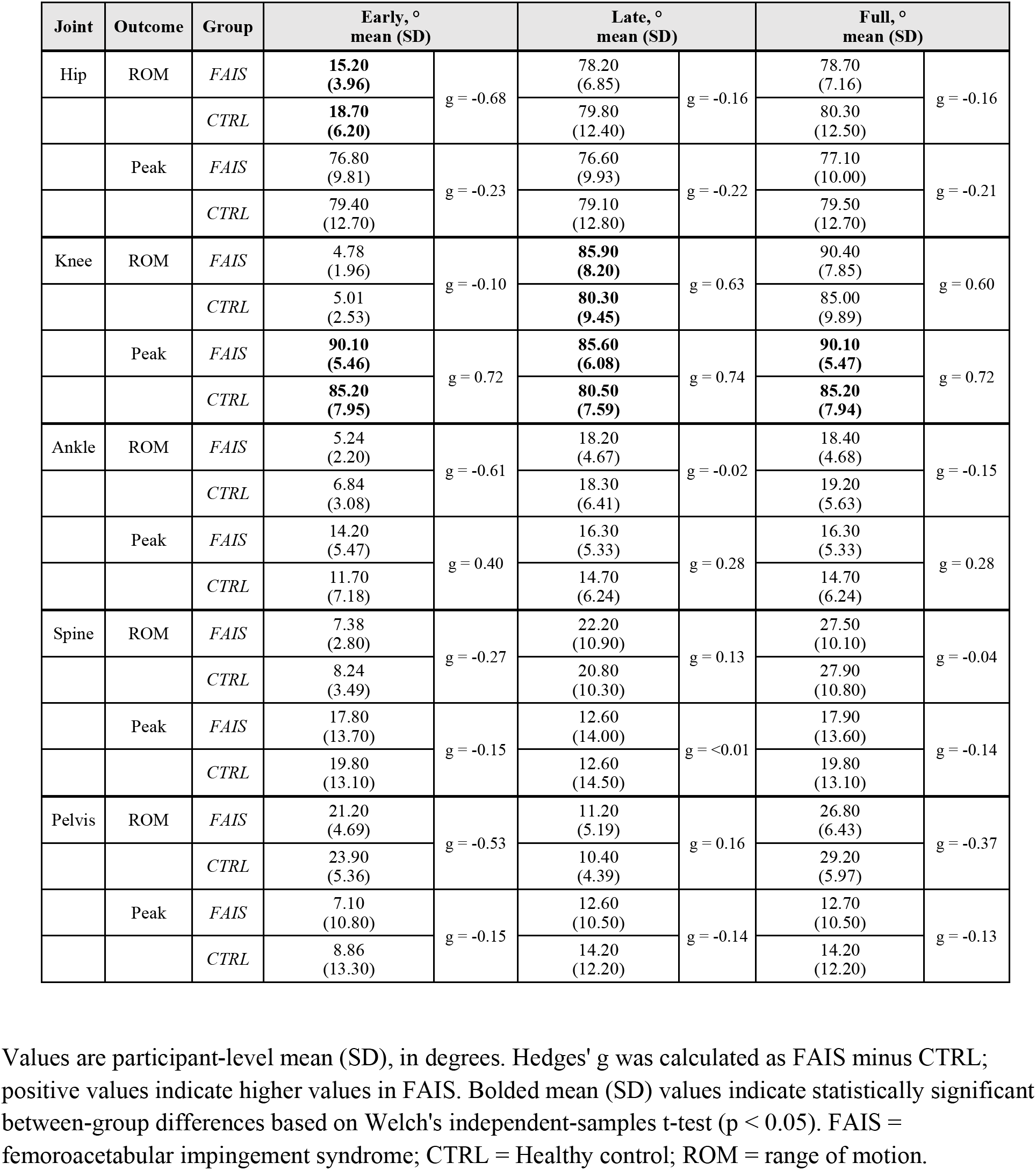
Sagittal-plane joint kinematics by group and sit-to-stand phase.

| Joint | Outcome | Group | Early, °<br>mean (SD) |  | Late, °<br>mean (SD) |  | Full, °<br>mean (SD) |  |
| --- | --- | --- | --- | --- | --- | --- | --- | --- |
| Hip | ROM | *FAIS* | **15.20**<br>**(3.96)** | g = -0.68 | 78.20<br>(6.85) | g = -0.16 | 78.70<br>(7.16) | g = -0.16 |
|  |  | *CTRL* | **18.70**<br>**(6.20)** |  | 79.80<br>(12.40) |  | 80.30<br>(12.50) |  |
|  | Peak | *FAIS* | 76.80<br>(9.81) | g = -0.23 | 76.60<br>(9.93) | g = -0.22 | 77.10<br>(10.00) | g = -0.21 |
|  |  | *CTRL* | 79.40<br>(12.70) |  | 79.10<br>(12.80) |  | 79.50<br>(12.70) |  |
| Knee | ROM | *FAIS* | 4.78<br>(1.96) | g = -0.10 | **85.90**<br>**(8.20)** | g = 0.63 | 90.40<br>(7.85) | g = 0.60 |
|  |  | *CTRL* | 5.01<br>(2.53) |  | **80.30**<br>**(9.45)** |  | 85.00<br>(9.89) |  |
|  | Peak | *FAIS* | **90.10**<br>**(5.46)** | g = 0.72 | **85.60**<br>**(6.08)** | g = 0.74 | **90.10**<br>**(5.47)** | g = 0.72 |
|  |  | *CTRL* | **85.20**<br>**(7.95)** |  | **80.50**<br>**(7.59)** |  | **85.20**<br>**(7.94)** |  |
| Ankle | ROM | *FAIS* | 5.24<br>(2.20) | g = -0.61 | 18.20<br>(4.67) | g = -0.02 | 18.40<br>(4.68) | g = -0.15 |
|  |  | *CTRL* | 6.84<br>(3.08) |  | 18.30<br>(6.41) |  | 19.20<br>(5.63) |  |
|  | Peak | *FAIS* | 14.20<br>(5.47) | g = 0.40 | 16.30<br>(5.33) | g = 0.28 | 16.30<br>(5.33) | g = 0.28 |
|  |  | *CTRL* | 11.70<br>(7.18) |  | 14.70<br>(6.24) |  | 14.70<br>(6.24) |  |
| Spine | ROM | *FAIS* | 7.38<br>(2.80) | g = -0.27 | 22.20<br>(10.90) | g = 0.13 | 27.50<br>(10.10) | g = -0.04 |
|  |  | *CTRL* | 8.24<br>(3.49) |  | 20.80<br>(10.30) |  | 27.90<br>(10.80) |  |
|  | Peak | *FAIS* | 17.80<br>(13.70) | g = -0.15 | 12.60<br>(14.00) | g = <0.01 | 17.90<br>(13.60) | g = -0.14 |
|  |  | *CTRL* | 19.80<br>(13.10) |  | 12.60<br>(14.50) |  | 19.80<br>(13.10) |  |
| Pelvis | ROM | *FAIS* | 21.20<br>(4.69) | g = -0.53 | 11.20<br>(5.19) | g = 0.16 | 26.80<br>(6.43) | g = -0.37 |
|  |  | *CTRL* | 23.90<br>(5.36) |  | 10.40<br>(4.39) |  | 29.20<br>(5.97) |  |
|  | Peak | *FAIS* | 7.10<br>(10.80) | g = -0.15 | 12.60<br>(10.50) | g = -0.14 | 12.70<br>(10.50) | g = -0.13 |
|  |  | *CTRL* | 8.86<br>(13.30) |  | 14.20<br>(12.20) |  | 14.20<br>(12.20) |  |

Values are participant-level mean (SD), in degrees. Hedges' g was calculated as FAIS minus CTRL; positive values indicate higher values in FAIS. Bolded mean (SD) values indicate statistically significant between-group differences based on Welch's independent-samples t-test ( $p < 0.05$ ). FAIS = femoroacetabular impingement syndrome; CTRL = Healthy control; ROM = range of motion.

### 3.3. Group differences in coordination variability

For the spine-hip coupling, a group*phase interaction (p = 0.001, ηₚ²= 0.252) was observed. Bonferroni-adjusted follow-up Welch’s t-test comparisons indicated a more variable coordination strategy during the early phase in the FAI group (mean difference = 6.21°*, p* = 0.010, Cohen’s d = 0.915), but not during the late phase (mean difference = −0.926°, *p* = 0.537, Cohen’s d = 0.205) (Figure 4). No other group effects or interactions were observed for the spine-hip and hip-knee couples (Table 3). Main effects of phase were observed for both joint pairs (p < 0.05), indicating that coordination variability differs between early and late STS, independent of group.

**Figure 4.**
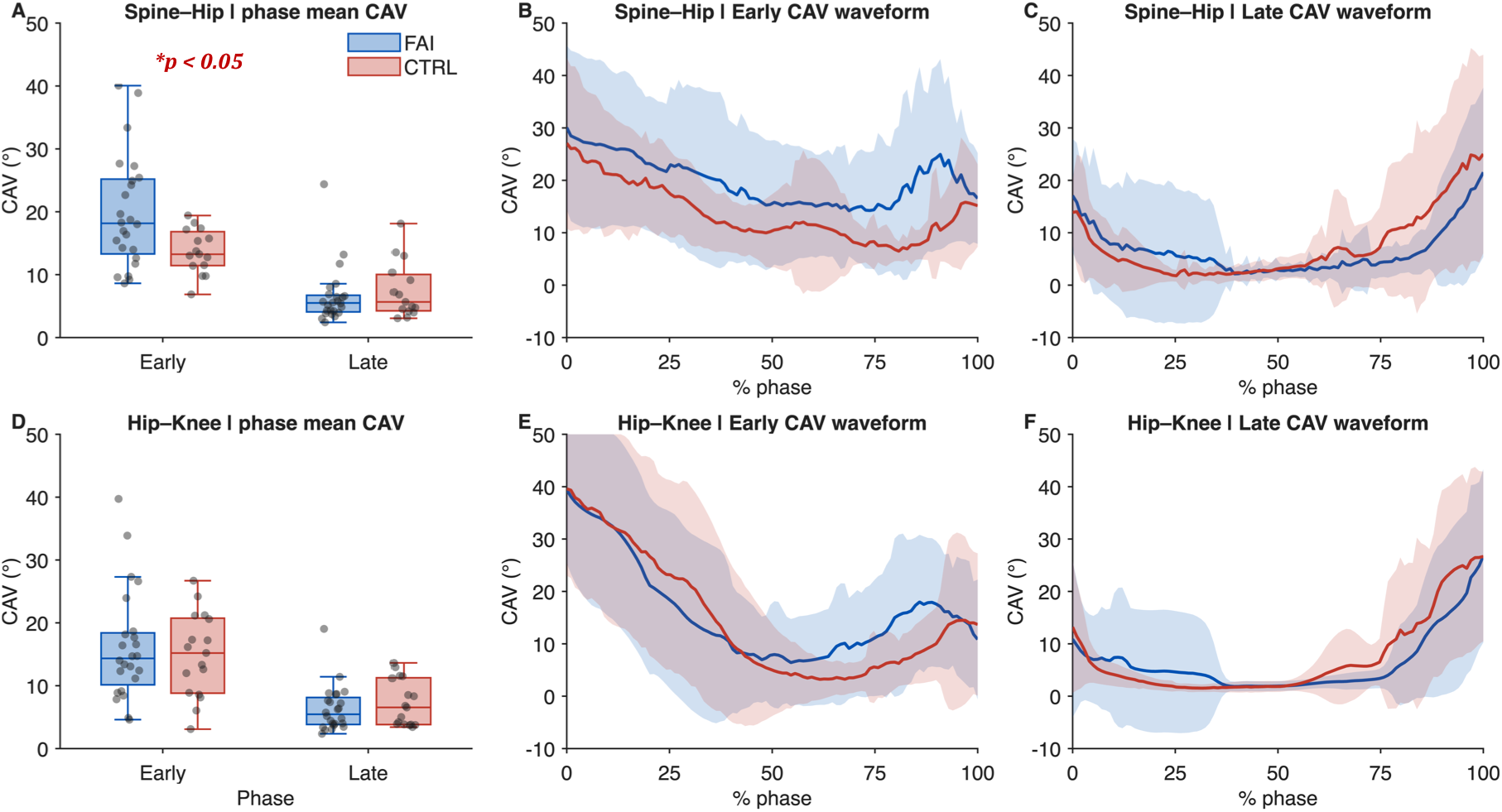
Phase-specific coupling-angle variability (CAV) for the femoroacetabular impingement (FAIS, blue) and Healthy control (CTRL, red) groups. Boxplots show participant-level mean CAV values for spine-hip (panel A) and hip-knee couplings (panel D), with individual data points overlaid. Adjacent waveforms (panels B,C,E,F) show group mean pointwise CAV across normalized early and late sit-to-stand phases, with shaded regions representing between-participant standard deviations. *denotes statistical significance.

**Table 3.** Coupling-angle variability by group and phase with mixed ANOVA results.

| Coupling | Group | Early CAV, °<br>mean (SD) | Late CAV, °<br>mean (SD) | Phase<br>p, ($\eta_p^2$) | Phase × Group<br>p, ($\eta_p^2$) | Group<br>p, ($\eta_p^2$) |
| --- | --- | --- | --- | --- | --- | --- |
| Spine–Hip | *FAIS* | 19.91 (8.92) | 6.57 (4.57) | **< 0.001**<br>**(0.717)** | **0.001 (0.252)** | 0.137<br>(0.059) |
|  | *CTRL* | 13.70 (3.54) | 7.49 (4.47) |  |  |  |
| Hip–Knee | *FAIS* | 16.19 (8.72) | 6.31 (3.62) | **< 0.001**<br>**(0.554)** | 0.379 (0.020) | 0.885<br>(0.001) |
|  | *CTRL* | 14.85 (6.64) | 7.22 (3.68) |  |  |  |

CAV = coupling-angle variability; FAIS = femoroacetabular impingement syndrome; CTRL = Healthy control; $\eta_p^2$ = partial eta squared. Values are participant-level mean (SD), in degrees. Phase refers to the within-participant effect of sit-to-stand phase (early and late); Group refers to the between-participant effect of group; Phase × Group refers to the interaction. Bolded values indicate statistical significance ( $p < 0.05$ ).
*[Figure removed in order to adhere with MedRxiv policy. Readers are encouraged to contact the corresponding author for access to this figure].*

### 3.4. Coordination strategies

Frequencies were interpreted descriptively as interdependent components of the overall coordination profile (Figure 5).

**Figure 5.**
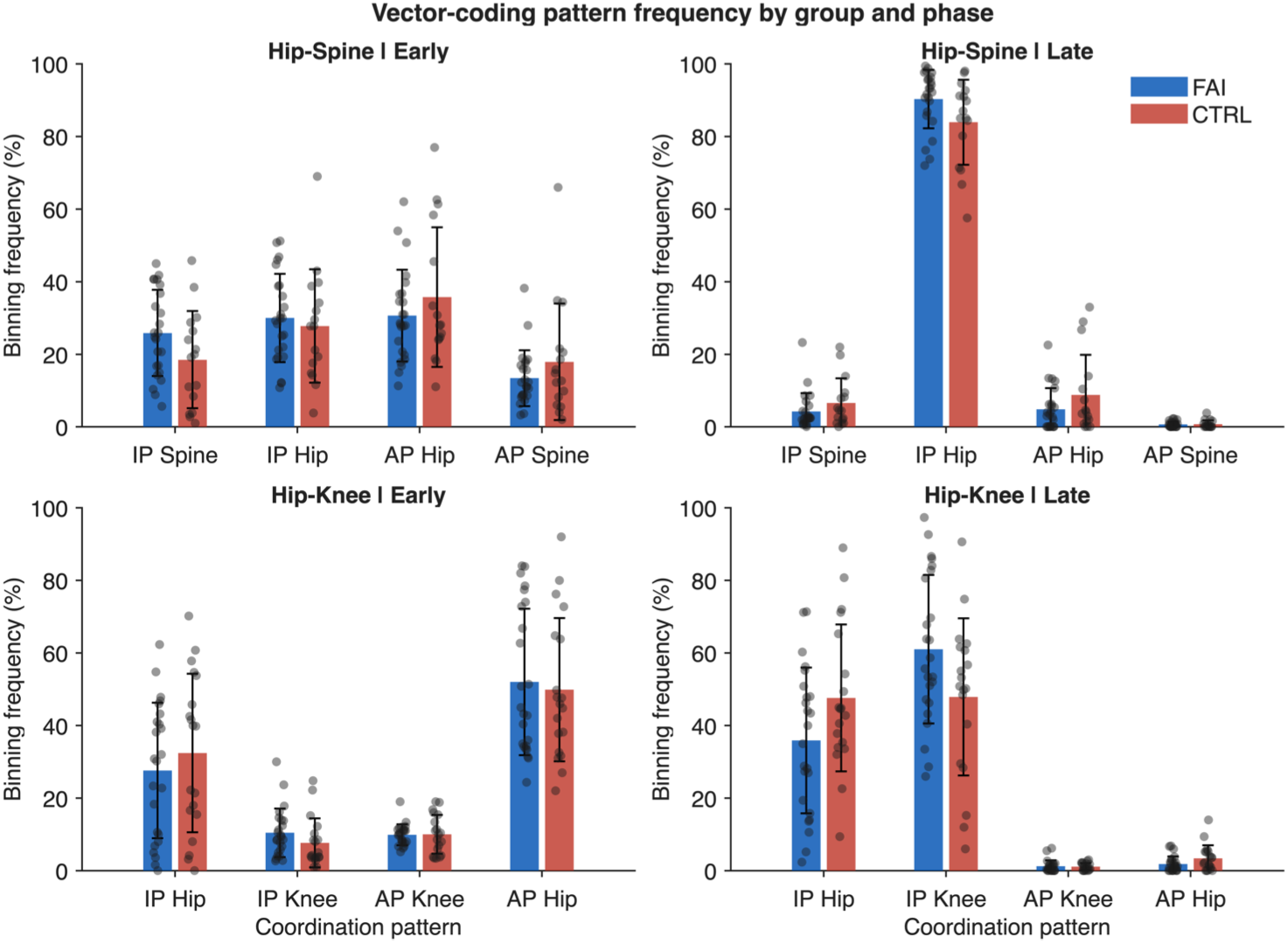
Vector-coding coordination-pattern frequencies by group and phase. Coordination-pattern frequencies are shown for spine–hip coupling (top row) and hip–knee coupling (bottom row) during the early and late phases in the femoroacetabular impingement (FAIS) and Healthy control (CTRL) groups. Bars represent group mean binning frequencies, error bars represent standard deviations, and grey points represent individual participant values. IP = in-phase; AP = anti-phase. The second label denotes the dominant segment within the coordination pattern.

During spine-hip early-phase STS, the FAI group showed a greater proportion of in-phase, spine-dominant coordination compared to healthy controls. In the late phase, both groups were predominantly in-phase, hip-dominant, with a modestly higher frequency in FAI.

During the early phase, healthy controls showed somewhat more in-phase, hip-dominant hip-knee coordination; otherwise, profiles were broadly similar. In the late phase, the FAI group showed relatively more in-phase, knee-dominant coordination, while healthy controls showed relatively more in-phase, hip-dominant coordination. The hip– knee late STS observation is the strongest pattern because it describes a redistribution between hip-and knee-dominant in-phase coordination.

## 4. DISCUSSION

This study compared sagittal plane kinematics and inter-joint coordination variability during STS in pre-operative individuals with cam-type FAI and healthy controls. SPM analyses revealed the FAI group and healthy controls had similar sagittal plane waveforms across the kinetic chain during STS, while secondary analyses revealed, on average, less hip ROM (early phase) and greater knee ROM (late phase) in FAI patients. Greater peak knee flexion across all phases of STS was observed in FAI patients; however, this likely reflected differences in initial seated posture. Individuals with cam-type FAI also demonstrated greater spine-hip, but not hip-knee, coordination variability during early STS. These findings partially supported our hypotheses and move beyond discrete biomechanical measures to provide insight into whole-body movement strategies in people with FAI.

### 4.1. STS kinematics were broadly similar, with subtle phase-specific differences

Compared to healthy controls, FAI patients did not show statistically different hip, knee, ankle, pelvis, or spine kinematics via SPM analyses. The discrete analyses did show reduced hip ROM during early STS, greater peak knee flexion across all phases, and greater knee ROM during late STS in the FAI group.

The most directly comparable study examining STS kinematics in people with and without FAI reported findings that were broadly consistent with ours, observing no between-group differences in peak sagittal plane hip, knee, pelvis, and trunk flexion angles (Spiker et al., 2022). Our discrete analyses, however, identified greater peak knee flexion in FAI patients. The consistency of this difference across STS phases, together with visual inspection of the group mean waveforms suggests that it may primarily reflect a difference in starting position rather than a distinct movement adaptation (Figure 3). Post hoc Welch’s t-tests comparing group differences in the mean knee flexion angle across the first 5 frames of the STS task supported this notion (mean difference = 4.94°; 95% CI = 0.54 to 9.35; p = 0.029), while no other differences in starting position were identified (Supplemental Table 1). Thus, despite efforts to standardize starting the STS position, the symptomatic limb of the FAI group was positioned, on average, in greater knee flexion at movement onset. Similarly, the greater knee ROM during late STS may also reflect the FAI group starting the phase in more knee flexion and extending towards a comparable standing position. It remains unclear whether this difference reflected a pain-related strategy, habitual seated posture, or residual variability in the starting position.

To account for differences in initial posture, the SPM analyses were referenced to the initial seated position. The lack of significant hip, spine and pelvic kinematic differences during STS in the SPM analyses was somewhat unexpected. Nevertheless, visual inspection of our group mean kinematic waveforms and SPM{t} trajectories (Figure 3, Supplemental figure 2), suggested small directional differences near seat-off, including smaller excursions towards anterior pelvic tilt and hip flexion, and greater spinal flexion excursion in the FAI group. Together, with the reduced early-phase hip ROM identified in the discrete analysis, these descriptive patterns raise the hypothesis that some people with FAI may redistribute sagittal plane motion away from the hip and towards the spine during the transition towards seat-off. More broadly, radiographic studies in hip pathology have shown that sagittal-plane deep-seated postures can be redistributed across the hip–pelvis– spine chain towards greater posterior pelvic tilt and lumbar flexion when hip motion is limited (Verhaegen et al., 2026). However, the population, task, and measurement approach differed from our present study. Future studies using imaging-based measures of spinopelvic posture (e.g., EOS x-ray) may help identify subtle differences not captured by marker-based kinematics.

Reduced pelvic mobility has been reported during deep-squatting in cam-type FAI cohorts (Bagwell et al., 2016; Catelli et al., 2018; Lamontagne et al., 2009; Ng et al., 2015). Interestingly, the direction of apparent pelvic motion in the present study (i.e., less excursion towards anterior tilt) differed from that commonly described during deep flexion (i.e., reduced posterior pelvic tilting). Less excursion towards anterior tilt may reflect the distinct mechanical demands of STS – which requires forward progression of the body’s centre of mass through varying contribution from the spine, pelvis, and hips – rather than a more absolute posteriorly tilted pelvis. We can speculate that these subtle differences (reduced excursion into hip flexion and anterior pelvic tilting, alongside greater spinal flexion) hint at a redistribution of sagittal-plane motion towards the spine; however, because the hip, pelvis, and spine SPM trajectories remained below the critical threshold, the proposed interpretation should be considered hypothesis-generating. More demanding tasks, such as single-leg squat, may be more likely to reveal consistent movement differences between people with and without FAI (Luijendijk et al., 2026).

### 4.2. Greater spine-hip coordination variability in FAI patients during early-phase STS

Compared to healthy controls, people with FAI demonstrated greater between-trial coordination variability at spine-hip, but not hip-knee, joint pairs during early STS. In related work, Hao et al., used vector coding to compare hip flexion-adduction coordination variability in people with and without FAI during a single-leg squat, reporting no between-group differences (Hao et al., 2026). Importantly, although this differed from our findings, Hao et al., examined multiplanar coordination within the hip, rather than a spine-hip coupling. Similar work in FAI reported altered coordination via smaller pelvic-femoral ratios (i.e., less relative pelvic contribution) during tasks such as squatting and stepping (Bagwell and Powers, 2019); however, the between-trial variability of this strategy was not explored. Otherwise, greater coordination variability has also been reported during early-phase STS in people with low back pain, at the lower lumbar-hip couple (Ippersiel et al., 2018), although different clinical populations complicate comparisons with our work. Few comparable studies exist, which makes situating our findings difficult.

Greater spine-hip coordination variability in early-phase STS could have multiple interpretations. First, this behavior might indicate that variability exceeded a functional range, reflecting less stable or robust neuromuscular control and potentially exposing tissues/structures to abnormal loading (Hamill et al., 2012; Stergiou and Decker, 2011). This interpretation is broadly aligned with a systematic review which reported a trend towards greater variability in people with lower-extremity musculoskeletal injury, although this review was neither specific to coordination variability nor STS (Baida et al., 2018). In FAI patients, such variability may become more apparent during the mechanically demanding transition towards seat-off, which requires substantial hip flexion. However, whether it alters joint loading or contributes to impingement remains speculative. Alternatively, greater spine-hip coordination variability may be an adaptive strategy to vary/redistribute load away from pain sensitive structures (Hamill et al., 2012; Hodges and Tucker, 2011). While hip joint contact forces have not been quantified in FAI patients during STS, modelling studies have demonstrated lower hip contact forces during key aspects of gait and squatting (Catelli et al., 2021; Ng et al., 2018; Savage et al., 2025). At face value, lower hip contact forces may be consistent with a protective unloading mechanism; however, Savage et al., reported that people with FAI walked with lower magnitude contact forces that were constrained to smaller regions of the acetabulum and femoral head (Savage et al., 2022), a pattern that may contribute to cartilage maladaptation. Thus, we cannot say whether greater spine-hip coordination variability in FAI patients is adaptive or maladaptive; studying coordination alongside joint and regional tissue loading may help answer this question.

Despite greater spine-hip coordination variability in FAI, no group differences were identified in the hip-knee joint pair. However, the cam-type FAI group demonstrated relatively more in-phase, knee-dominant coordination during the late phase of STS, whereas the healthy group exhibited relatively more in-phase, hip-dominant coordination (Figure 4). Although this late-phase finding is a qualitative description, it may suggest a relative shift from hip-toward knee-dominant motion during rising.

### 4.3. Limitations

This work has certain limitations. First, we used a cross-sectional study design. Thus, causal relationships between cam-type FAI and the observed biomechanics cannot be established. Additionally, our sample was predominately male with unbalanced group sizes. While cam-type FAI is more common in males, this nevertheless limits the generalizability of our findings for females. Additionally, the older age in our FAI sample may have impacted our findings. Further, the self-selected comfortable speed and STS motion itself may not have been challenging enough to elicit the full range of compensatory strategies in FAI patients. For context, Bagwell et al., reported peak hip flexion during squat in cam-type FAI patients reached 106.6° (Bagwell et al., 2016), while our peak hip flexion during STS was 77.1°; thus, perhaps STS does not challenge the hip sufficiently to elicit adaptation. Also, the CAV was calculated using a maximum of 5 trials, which may limit the reliability of our variability estimates. Despite our efforts to standardize the STS starting position, it appears the FAI group had more knee flexion on the affected side. To account for this, we referenced our SPM analyses to the starting position; however, this may have influenced our results. Last, compared with imaging-based subject-specific models, the use of generic marker-based models has been shown to introduce systematic offsets in sagittal plane hip and pelvic angle estimates in tasks involving substantial hip flexion. While our SPM analyses were offset to seated posture which helps mitigate this issue, the use of generic models may have influenced our estimates of absolute joint angles and peak angles measures (Al Otti et al., 2024).

## 5. CONCLUSION

Compared to healthy controls, people with FAI had greater spine-hip coordination variability and reduced hip ROM during early-phase STS, despite broadly similar kinematic waveforms. This suggests that movement adaptations during STS may be reflected more strongly in inter-joint coordination than in changes in individual joint kinematics.

## Data Availability

All data produced in the present study are available upon reasonable request to the authors

## Acknowledgements

The authors would like to thank Adam Allam, Hallie Maron, and Charlie Tierney for their help with data collection and processing. The authors also thank the research staff of the Division of Orthopaedic Surgery, University of Ottawa, for help in patient recruitment and data collection. This work was supported by the Canadian Institutes of Health Research (CIHR) under Grant Number 400348.

## Use of artificial intelligence disclaimer

Chat-GPT (OpenAI) was used to help with: (i) phrasing, grammar, and writing clarity in specific manuscript sections; (ii) to assist with identifying potentially relevant literature; and (iii) troubleshooting when developing MATLAB code. All literature, citations, code, analyses, and manuscript content were reviewed and verified. The authors take full responsibility for the accuracy of the final manuscript.

