## Supplemental Figure 1 for "Sit-to-Stand Adaptations in Pre-Operative Cam-Type Femoroacetabular Impingement: A Kinematic and Coordination Analysis"

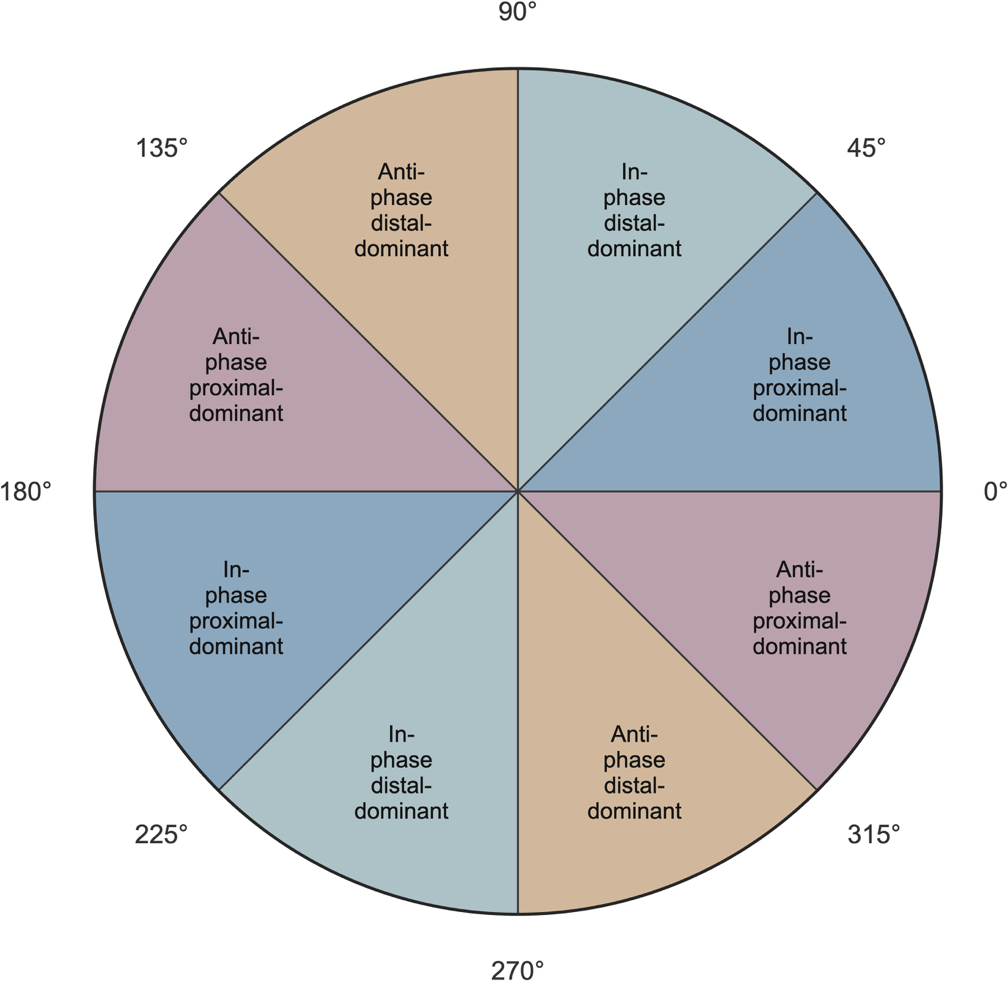

**Supplemental Figure 1. Vector-coding coordination-pattern classification.** Coupling angles were classified as in-phase proximal-dominant (0° ≤ CA < 45° and 180° ≤ CA < 225°), in-phase distal-dominant (45° ≤ CA < 90° and 225° ≤ CA < 270°), anti-phase distal-dominant (90° ≤ CA < 135° and 270° ≤ CA < 315°), or anti-phase proximal-dominant (135° ≤ CA < 180° and 315° ≤ CA < 360°). The proximal and distal variables occupied the horizontal and vertical axes, respectively.

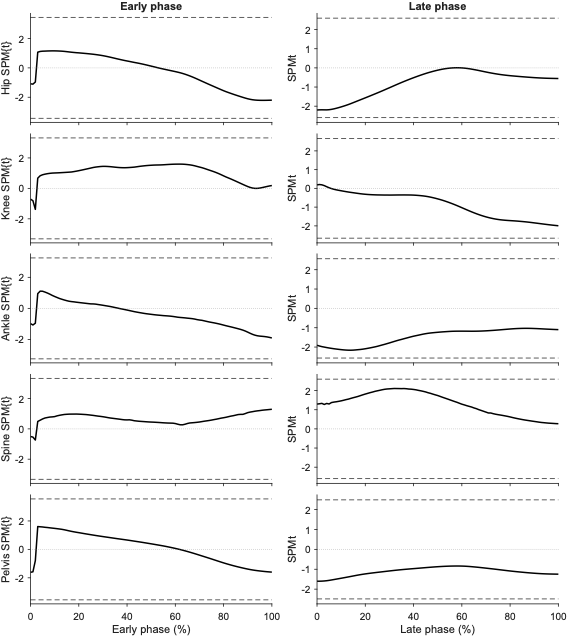

**Supplementary Figure 2. Statistical parametric mapping results for offset-referenced sagittal-plane kinematics during sit-to-stand.** SPM{t} trajectories are shown for the early and late phases for the hip, knee, ankle, spine, and pelvis. Solid black lines represent the SPM{t} statistic comparing individuals with femoroacetabular impingement syndrome (FAIS) with Healthy controls (CTRL). Dashed horizontal lines indicate the critical SPM threshold. Grey vertical shading identifies regions where statistically significant between-group differences were detected.

**Supplemental Table 1. Starting sagittal-plane joint and segment angles by group**

| **Joint /** **segment** | **Group** | **Starting position, °** **mean (SD)** | **Mean difference, °** **(95% CI)** | **Welch** **p value** |
| --- | --- | --- | --- | --- |
| Hip | *FAI* | 61.69 (10.32) | 0.57 (-5.78 to 6.91) | 0.857 |
|  | *CTRL* | 61.13 (10.13) |  |  |
| Knee | *FAI* | **89.53** **(5.90)** | 4.94 (0.54 to 9.35) | **0.029** |
|  | *CTRL* | **84.59** **(7.84)** |  |  |
| Ankle | *FAI* | 9.35 (6.07) | 3.53 (-1.40 to 8.45) | 0.152 |
|  | *CTRL* | 5.83 (7.28) |  |  |
| Spine | *FAI* | 13.50 (12.46) | -1.60 (-9.23 to 6.04) | 0.674 |
|  | *CTRL* | 15.10 (11.45) |  |  |
| Pelvis | *FAI* | -13.96 (10.74) | 0.15 (-7.03 to 7.34) | 0.965 |
|  | *CTRL* | -14.12 (10.65) |  |  |

Values represent the mean angle across the first five normalized samples of early STS. Mean differences are calculated as FAI (femoroacetabular impingement) minus CTRL (healthy control). Bolded values denote statistical significance (p < 0.05)
